# Autonomic Nervous System Markers of Music-Elicited Analgesia in People with Fibromyalgia: A Double-Blind Randomized Pilot Study

**DOI:** 10.1101/2022.05.25.22275605

**Authors:** Rebecca J. Lepping, Miranda L. McMillan, Andrea L. Chadwick, Zaid M. Mansour, Laura E. Martin, Kathleen M. Gustafson

## Abstract

**Purpose:** To investigate the feasibility of using music listening by adults with fibromyalgia (FM) as a potential tool for reducing pain sensitivity.

**Patients and methods:** We report results from a double-blind two-arm parallel randomized pilot study (NCT04059042) in 9 participants with FM. Pain tolerance and threshold were measured objectively using quantitative sensory tests; autonomic nervous system (ANS) reactivity was measured with electrocardiogram. Participants were randomized to listen to instrumental Western Classical music or a nature sound control to test whether music listening elicits greater analgesic effects over simple auditory distraction. Participants also completed separate control testing with no sound that was counterbalanced between participants.

**Results:** Participants were randomized 1:1 to music or nature sounds (4 Music, 5 Nature). Although the groups were not different on FM scores, the Music group had marginally worse pain summation (p=.09) and tolerance (p=.13). The Nature group had higher Anxiety scores (p<.05). Outcome measures showed a significant Group by Session interaction for pain tolerance (p<.05) revealing that the Nature group had greater pain reduction during audio compared to silence, while the Music group had no difference between sessions. No significant effects were observed for ANS testing. Within the Music group, there was a trend of vagal response increase from baseline to music listening, but it did not reach statistical significance; this pattern was not observed in the Nature group.

**Conclusion:** Auditory listening significantly altered pain responses. There may be greater vagal response to music versus nature sounds, however, results could be due to group differences in pain and anxiety. This line of study will help determine whether music could be prophylactic for people with FM when acute pain is expected.

## 1 Introduction

The term “centralized pain” describes any central nervous system (CNS) dysfunction or pathology that may contribute to the development or maintenance of chronic pain.(1-3) There is a growing appreciation of the role of CNS augmentation of pain processing in many chronic pain conditions.(2, 4) A hallmark of the centralized pain phenotype is the presence of hyperalgesia and reduced or absent endogenous analgesia.(5-7) Data from quantitative sensory testing (QST) studies suggest a wide, bell-shaped distribution in pain sensitivity across the general population. Most individuals with centralized pain fall on the right side of this curve and have QST findings consistent with hypersensitivity (hyperalgesia and allodynia).(1, 8-14) QST evidence of widespread hypersensitivity is consistently observed in many chronic pain conditions, including FM, irritable bowel syndrome, tension headache, low back pain, temporomandibular joint disorder, interstitial cystitis, and vulvodynia.(15-25) In this study, we used QST to objectively measure pain sensitivity while listening to music compared to listening to nature sounds, in patients with FM.

Music has been previously shown to influence parameters of the autonomic nervous system associated with anxiety, (26) such as slowing heart rate and respiration.(27) Music listening can also reduce acute pain during surgery,(28) post-operative recovery,(29) orthodontic procedures,(30) and during thermic pain induction in healthy participants.(31) The *subjective* analgesic, anxiolytic, and antidepressant effects of music for people with chronic pain were recently confirmed in a meta-analysis.(32) However, the impact of music listening on objective measures of pain sensitivity in patients with chronic pain has not yet been described. The goal of this pilot study was to understand possible analgesic effects of music listening on objective measures of pain sensitivity in patients with fibromyalgia (FM).

The analgesic effect of music is thought to occur through several mechanisms: Contextual, Cognitive, Emotional, and Physiological.(33) First, music provides a predictable *context* that can increase the listeners’ sense of control. This is further enhanced if the music is familiar, as this can bring in other effects that are not related to aspects of music specifically, such as setting up expectations and heightening nostalgia. Studies have shown the greatest analgesic effects when music is selected by participants. Second, similar to other types of stimulation such as reading or listening to nature sounds, (34) music can serve as a *cognitive* distraction and take attention away from the painful stimulus. Third, music is a powerful inducer of *emotion*.(35, 36) Music that is positive, liked by the listener, and low on arousal has the strongest analgesic effect.(31) Finally, music listening interventions and music therapy have also been shown to reduce anxiety and depression.(37, 38) The anxiolytic effect may be due to the *physiological* effect of music on the parasympathetic nervous system, increasing the vagal response and reducing heart rate and respiration rate.(26) Music also has effects on the brain directly, causing the release of endogenous opioids and dopamine and activating areas of the descending pain modulatory system.(39, 40) The specific musical characteristics that yield the greatest analgesic effects are difficult to pinpoint, as there is not a standard for reporting. Meta-analyses have revealed that music with 60-80 beats per minute, in a major key and without lyrics or percussion have the largest effects.(41)

Previous studies in patients with FM have shown that patients have reduced self-reported pain and increased mobility after even a short, 10-minute music listening intervention. After listening to music of their choice, participants were faster in a standard mobility assessment, the timed-up-and-go task.(42) A second study using resting state functional magnetic resonance imaging confirmed the impact of a 5-minute music listening intervention on the centralized descending pain modulatory system (DPMS), identified as changes in functional connectivity between regions of the DPMS that positively correlated with changes in pain scores.(43) Our study is the first to investigate whether objectively measured pain sensitivity is reduced by music listening.

The goal of the current study was to identify whether music listening has a promising analgesic effect during pain threshold and tolerance testing for patients with FM that supersedes any effect of auditory distraction. We hypothesized that because the nature listening condition provides a distraction from pain sensations, and may also provide some of the same Contextual, Cognitive, Emotional, and Physiological impacts as Music, both listening conditions (Music and Nature) would reduce pain sensitivity compared to testing during Silence. However, as noted previously, the Emotional and Physiological impacts are anticipated to be stronger in Music due in part to temporal structure and expectancy building, we therefore hypothesized that Music listening would reduce pain sensitivity compared to Nature sounds. We further hypothesized that Music would increase vagal input to the autonomic nervous system, decreasing heart rate and increasing heart rate variability compared to both Silence and Nature sounds, and that analgesic responsiveness would be moderated by symptoms of FM, anxiety, and depression.

## 2 Material and methods

### 2.1 Trial Design

This was a single center, two-arm parallel double-blind randomized controlled pilot study conducted in the United States (ClinicalTrials.gov NCT04059042). Participants with FM underwent two testing sessions, one week apart: testing as usual with no-sound (Silence), and testing while listening to instrumental Western Classical music, or nature sound control (Audio). Participants were randomized 1:1 to the two arms (Music or Nature sounds), counterbalanced for session order. Study data were collected and managed and randomization was implemented using REDCap electronic data capture tools (44, 45) using a computer-generated random order for the four possible session orders (Music/Silence, Silence/Music, Nature/Silence, Silence/Nature), and was stratified by gender.

#### 2.1.1 Changes to Trial Design

The study was stopped early due to the COVID-19 pandemic.

### 2.2 Participants

Participants with a diagnosis of FM were recruited from pain clinics at a large Midwestern US university medical center and by word of mouth. Eligible participants were 18 years or older, able to read and speak English, willing to refrain from alcohol, nicotine, and physical activity or exercise on the day of testing, and on a stable dose of adjunctive pain medications, including tricyclic antidepressants, serotonin-norepinephrine reuptake inhibitors, and gabapentinoids. Participants were excluded if they were not able to provide written consent, were pregnant, had peripheral neuropathy in the upper extremities, had severe physical impairment or co-morbid medical conditions such as blindness, deafness, paraplegia, cancer, autoimmune disorder, liver failure or cirrhosis, hepatitis, cardiovascular disease, illicit drug or opioid abuse, or average daily opioid dosing of >15 mg oral morphine equivalents (e.g., > two 5 mg oxycodone tablets/day or > three 5 mg hydrocodone tablets/day). Conversions were made based on well-accepted conversion tools.(46, 47)

#### 2.2.1 Study Setting

The study was conducted at a research laboratory within the medical center campus. Data were collected with participants seated in a small, quiet room across a small table from the researcher.

### 2.3 Procedures (Interventions)

Participants in both arms had QST and electrocardiogram (ECG) testing on two separate days, one week apart: baseline (Testing as Usual, Silence) and auditory listening (Music or Nature sounds), counterbalanced across participants. After completing informed consent, participants were fitted with ECG electrodes and were given instructions about the procedures. Participants were given noise-cancelling headphones to wear during all testing procedures. The researcher wore ear plugs to remain blinded to what the participant was hearing and communicated with the participant through written instructions and gestures for the remainder of the test. After the electrodes and headphones were in place, the researcher left the room and baseline ECG was recorded for five minutes while participants sat quietly. The researcher returned to the room, started the specified audio track, and then left the room for 10 minutes while participants sat quietly listening to the track. The researcher then returned to the room for QST testing while the participant continued to listen to the audio track. Written instruction reminders were provided to participants before each task. At the end of the first day of testing, participants completed surveys electronically on a laptop through REDCap. (44, 45) After completing all procedures on the second day of testing, participants were given $100 for their time.

#### 2.3.1 Music and Sound Delivery

Auditory stimuli were presented using a digital music player and noise-cancelling headphones. Four audio tracks were identified by number only, and the researcher was blinded to the contents of each tract. One track was Music, one was Nature sounds, and two were Silence. The randomization procedure indicated to the researcher which track (1-4) should be used for the testing session. Each track began with instructions to the participant indicating what they would hear during testing, and that they should continue to wear the headphones even if the track is Silence so that the researcher would not know what they were hearing.

#### 2.3.2 Music Characteristics

The musical selections consisted of professional recordings of instrumental Western Classical music selected by the researcher (Supplementary Table 1). All participants heard the same pieces in the same order. Instrumentation ranged from piano solo to full orchestra, but all were without lyrics or heavy percussion. Pitch ranged across pieces but was standard across participants and not controlled by either the participant or the researcher. Tempo for all pieces was slow (∼60 beats per minute). The pieces were in either major keys or minor keys, but all consisted primarily of consonant harmonies and sustained melodic phrases. Participants were allowed to control the volume to their individual comfort level.

**Table 1.** Participant demographic variables by audio group assignment (Music, Nature)

| | Music Group<br>(*n* = 4) | Nature Group<br>(*n* = 5) | *t* / $X^2$ | *p* | 95% *CI* |
| --- | --- | --- | --- | --- | --- |
| Age (years) (*M* (*SD*)) | 49.18 (13.86) | 40.28 (9.93) | 1.13 | .30 | [-9.78, 27.59] |
| Dominant Hand, Right (*n* (%)) | 4 (100%) | 5 (100%) | -- | -- | -- |
| Gender, Female | 4 (100%) | 5 (100%) | -- | -- | -- |
| Sex Assigned at Birth, Female | 4 (100%) | 4 (80%) | 0.90 | .34 | -- |
| Race, White | 4 (100%) | 5 (100%) | -- | -- | -- |
| Ethnicity: Not Hispanic or Latinx | 2 (50%) | 5 (100%) | 3.21 | .20 | -- |
| Hispanic or Latinx | 1 (25%) | 0 (0%) |  |  |  |
| Other/Unknown/No Response | 1 (25%) | 0 (0%) |  |  |  |
| Relationship Status: Married | 2 (50%) | 4 (80%) | 3.60 | .31 | -- |
| Never Married | 1 (25%) | 0 (0%) |  |  |  |
| Divorced or Separated | 1 (25%) | 1 (20%) |  |  |  |
| Education: High School/ GED | 0 (0%) | 0 (0%) | 3.94 | .27 | -- |
| Some College | 1 (25%) | 1 (20%) |  |  |  |
| Technical/ Associate's Degree | 0 (0%) | 1 (20%) |  |  |  |
| Bachelor's Degree | 0 (0%) | 2 (40%) |  |  |  |
| Advanced/ Professional Degree | 3 (75%) | 1 (20%) |  |  |  |

**Notes:** Continuous measures were assessed with independent samples *t* tests; categorical variables were assessed with Chi-square tests.
**Abbreviations:** CI, confidence interval; M, mean; SD, standard deviation; GED, general education development.

#### 2.3.3 Active Control

Professional recordings of nature sounds (including forest, river, and wind sounds and birdsong) selected by the researcher without added music were used as the active control condition (Supplementary Table 1). All participants heard the same recording. This active control condition allowed for non-musical analgesic effects, such as distraction, to be controlled in the experimental design. Participants were allowed to control the volume to their individual comfort level.

### 2.4 Outcomes

#### 2.4.1 Autonomic Nervous System Activity (ECG)

Participants’ ECG were recorded using three standard snap-on ECG electrodes with Biopac MP150 and Acqknowledge 4.3 software (Goleta, CA). ECG electrodes were placed under the collar bone and below the rib cage on the opposite side, with a ground electrode placed on the abdomen near the navel. The time of each condition (Baseline, Listening only, and Pain while listening) was recorded by the investigator with a mark in the Acqknowledge recording. The ECG data were uploaded to Kubios software (Kuopio, Finland) for analysis. Summary metrics of heart rate and variability during each condition were corrected for within-session baseline levels and compared between conditions (Listening only vs. Pain while listening) and between auditory groups (Music vs. Nature sounds).

#### 2.4.2 Patient Characteristics Related to Pain and Music

##### Demographics

Participants completed a demographics questionnaire that included questions on participant sex, gender, age, race, ethnicity, marital status, education level, body mass index, and current medications.

##### Fibromyalgia-ness (FMness)

FMness is a measure of pain and co-morbid symptom extensiveness and severity, calculated by combining the scores from the Widespread Pain Index with the Symptom Severity Scale from the 2011 FM Survey(48) to derive a continuous metric purportedly indicative of the degree of CNS pain amplification present in a given individual.(49)

##### Clinical Pain Severity

Pain severity and functional interference due to pain were assessed using the Brief Pain Inventory (BPI). The BPI is validated for chronic, non-malignant forms of pain, and asks patients to rate their current pain intensity, as well as their worst, least and average pain in the 7 days (0-10 NRS) and has been recommended by IMMPACT as a measure of choice for the assessment of pain in clinical research.(50-52)

##### Fibromyalgia Functional Status

Current health and functional status in FM patients was measured using the Revised Fibromyalgia Impact Questionnaire (FIQR).(53) The FIQR measures physical functioning, work status, and overall well-being.

##### Depression and Anxiety

Mood symptoms were assessed with the static short forms for Depression and Anxiety, developed by the NIH roadmap initiative PROMIS.(54) The PROMIS measures have a standardized mean of 50, a standard deviation of 10, and a range of 1-100.

##### Music Experience

Participants rated their music listening habits (i.e. frequency, styles, reasons for listening, etc.) using the Brief Music Experience Questionnaire (MEQ)(55). The Brief MEQ is a 53 item self-report measure of music centrality in the respondent’s life, their musical aptitude, and experience with and reaction to music. Questions are rated using a 5-point Likert scale (1: very untrue; 5: very true), from which six summary scores are derived for Commitment to Music, Innovative Musical Aptitude, Social Uplift, Affective Reactions to Music, Positive Psychotropic Effects from Music, and Reactive Musical Behavior.

#### 2.4.3 Quantitative Sensory Testing (QST)

Pain testing was performed using the Multimodal Automated Sensory Testing (MAST) System, a computerized QST device developed at the University of Michigan, and currently being employed in several clinical trials, including the NIH MAPP Network. Two measures of QST were used in this study: mechanical pain sensitivity (MPS) and temporal summation (TS). MPS was assessed by applying discrete pressure stimuli to the thumbnail bed (Figure 1A). The MAST system delivered an ascending series of 5-s duration stimuli at 25-s intervals, beginning at 0.50 kg/cm2 and increasing in 0.50 kg/cm2 intervals up to tolerance or a maximum of 10 kg/cm2. Participants rated pain intensity after each stimulus on a 0 (no pain) – 100 (extreme pain) numerical rating scale (NRS). Pain threshold, the point at which participants rated >0 pain, and tolerance, the point at which participants rated >80 pain, were determined from this procedure. To measure TS, a 256 mN pinprick stimulus (MRC Systems, Heidelberg, Germany, Figure 1B) was applied once to the forearm or hand, followed by a train of 10 identical stimuli at a rate of 1 Hz. Following the single stimulus and the train of 10 stimuli, patients reported the pain intensity of the pinprick sensation using the 0-100 NRS. This procedure was repeated three times, and the mean pain rating of the three stimulus trains was divided by the mean pain rating of the single stimuli to calculate a wind-up ratio (WUR); a WUR >1 indicates temporal summation.^82^

**Figure 1.**
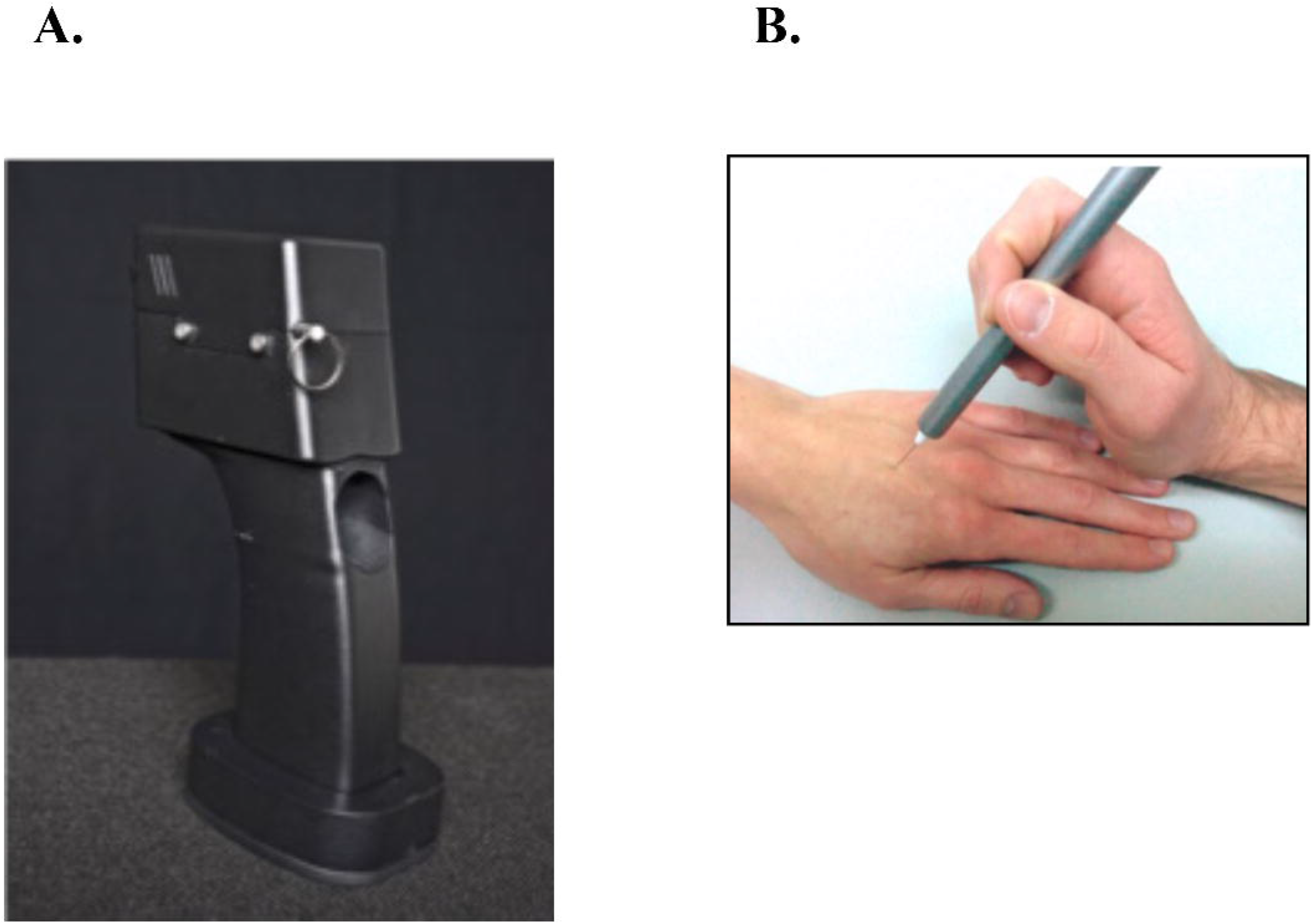
**A**. Multimodal Automated Sensory Testing (MAST) stimulator for assessing mechanical pain tolerance, and **B**. Pinprick stimulator used to assess temporal summation.

### 2.5 Sample Size

Ten White female participants with FM were enrolled in the study (Table 1). Centralized pain and nearly any chronic pain condition is 1.5 – 2 times more common in women than in men.(56) One person in the music group did not return for the second visit and was lost to follow-up. That person only received the Silence session and is not included in the analysis (Figure 2).

**Figure 2.**
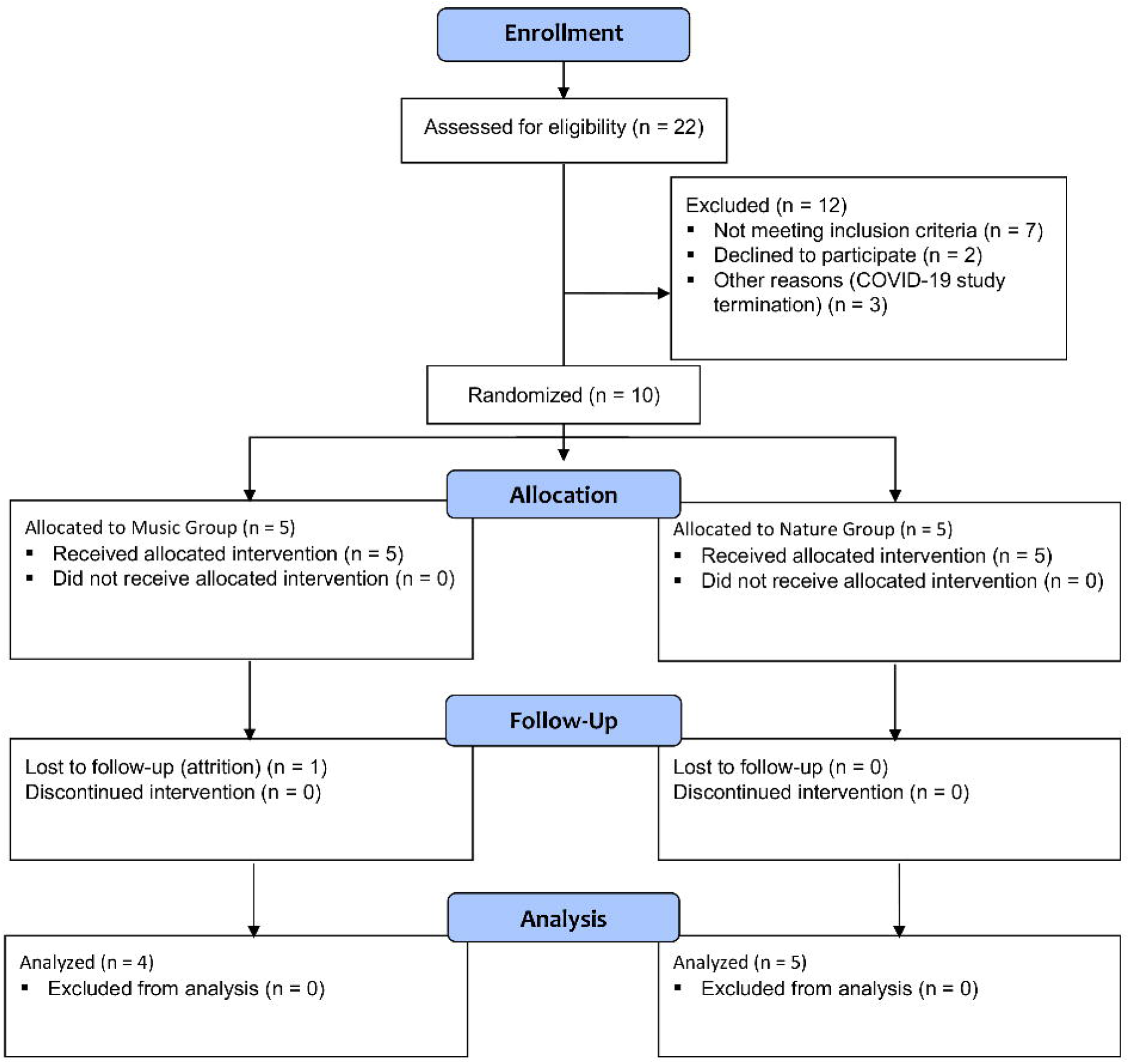
CONSORT 2010 Flow Diagram.

The intended sample size was 40 participants with FM based on power analysis, however, due to the COVID-19 pandemic, recruitment was stopped and only 10 participants took part in this study.

### 2.6 Randomization

#### 2.6.1 Randomization Sequence Generation

Randomization was defined using an online random number generator.

#### 2.6.2 Randomization Type

Patients were randomized 1:1 to Music or Active Control (Nature sounds), counterbalanced for session order with Silence. Randomization was implemented with the REDCap Randomization tool (44, 45) using a computer-generated random order for the four possible session orders (Music/Silence, Silence/Music, Nature/Silence, Silence/Nature), coded by track number only, and was stratified by gender.

#### 2.6.3 Randomization Allocation/Concealment Method and Implementation

Audio tracks for Music, Nature sounds, and two tracks for Silence were labelled with dummy codes (1-4) to blind the researcher collecting the data. The original audio tracks were given to a person outside the study team who renamed the files and placed the code into a sealed opaque envelope. The researcher selected the track by number assigned during randomization. Randomization was concealed from the researchers until final group analysis.

#### 2.6.4 Blinding

The study team were blinded throughout data collection and analysis. Sound cancelling headphones for the participant, earplugs for the researcher, and randomization of sessions were employed to blind the researcher to which audio track the participant was hearing and minimize bias.

#### 2.6.5 Similarity of Interventions

The participant wore headphones for all sessions, regardless of what they were hearing (Music, Nature sounds, or Silence). All sessions were conducted in the same way and lasted approximately the same amount of time.

### 2.7 Statistical Methods

Demographic characteristics and questionnaire measures were compared between the two Audio Groups using Student’s *t*-tests for continuous variables and Chi square (*Χ*^2^) for categorical variables. Outcome measures were assessed using repeated measures analysis of variance (ANOVA). Pain measures of mechanical pain tolerance and temporal summation, which were collected once per session, were assessed with a 2 (Audio Group: Music, Nature) X 2 (Session: Silence, Audio) repeated measures ANOVA. ANS measures of heart rate and heart rate variability (root mean square of successive differences, HRV) were assessed with a 2 (Audio Group: Music, Nature) X 2 (Session: Silence, Audio) X 2 (Condition: Listen, Pain) repeated measures ANOVA. Statistical significance was set at p < .05 for each test.

#### 2.7.1 Additional Analyses

To account for variability related to FMness, depression, and anxiety symptoms, a second set of ANOVAs was conducted including FMness, depression, and anxiety symptom as covariates. Further relationships were explored between the questionnaire measures and the magnitude of change in pain and ANS variables between sessions using Pearson’s correlations. Statistical significance was set at p<.05 for each test.

## 3 Results

### 3.1 Demographic and Questionnaire Measures

Group differences in demographic measures are presented in Table 1. The groups did not differ on age, gender, ethnicity, relationship status, or education level. Questionnaire measures are presented in Table 2. Participants in both groups were experiencing moderate FM, depression, and anxiety symptoms. They also reported low to moderate commitment to music and innovative musical aptitude, but reported moderate to high affective reactions to music, positive psychotropic effects from music and reactive musical behavior. The groups did not differ in FM symptom severity or music experience; however, they were significantly different on symptoms of anxiety, with participants in the Nature group experiencing higher anxiety than participants in the Music group.

**Table 2.** Participant reported clinical and musical experience variables by audio group assignment (Music, Nature)

|  | Music Group<br>(*n* = 4) | Nature Group<br>(*n* = 5) | *t* | *p* | 95% *CI* |
| --- | --- | --- | --- | --- | --- |
| WOLFE FMness (*M* (*SD*)) | 17.00 (4.76) | 13.00 (3.74) | 1.42 | .20 | [-2.68, 10.68] |
| BPI Worst 2 Average | 4.25 (2.63) | 4.20 (1.64) | .04 | .97 | [-3.32, 3.42] |
| FIQR Score | 45.50 (22.19) | 40.23 (15.78) | 0.42 | .69 | [-24.55, 35.08] |
| PROMIS: Depression | 45.98 (5.30) | 50.40 (5.79) | -1.18 | .28 | [-13.28, 4.43] |
| PROMIS: Anxiety | 52.65 (4.71) | 59.92 (3.33) | -2.72 | .03* | [-13.59, -0.95] |
| MEQ: Commitment to Music | 2.04 (0.92) | 1.60 (0.71) | 0.81 | .45 | [-0.84, 1.72] |
| MEQ: Innovative Musical Aptitude | 2.11 (0.63) | 2.17 (1.08) | -0.11 | .92 | [-1.51, 1.38] |
| MEQ: Social Uplift | 2.62 (0.48) | 2.70 (1.22) | -0.12 | .91 | [-1.62, 1.47] |
| MEQ: Affective Reactions to Music | 4.03 (0.84) | 4.40 (0.33) | -0.93 | .39 | [-1.33, 0.58] |
| MEQ: Positive Psychotropic Effects from Music | 3.53 (1.00) | 3.45 (0.58) | 0.15 | .88 | [-1.17, 1.33] |
| MEQ: Reactive Musical Behavior | 3.58 (0.57) | 4.00 (0.66) | -1.00 | .35 | [-1.40, 0.57] |

**Notes:** Continuous measures were assessed with independent samples *t* tests. \*Indicates significant group differences at *p* < .05.
**Abbreviations:** CI, confidence interval; M, mean; SD, standard deviation; FM, fibromyalgia; BPI, Brief Pain Inventory; FIQR, Fibromyalgia Impact Questionnaire – Revised; PROMIS, Patient-Reported Outcomes Measurement Information System; MEQ, Music Experience Questionnaire.

### 3.2 Pain Measures

In the ANOVA for temporal summation, the difference between a single stimulus and a series of stimuli, there was a significant interaction of Group by Session, with the Nature group showing lower temporal summation while listening to the audio compared to silence, while the Music group was not different between sessions. The main effect for Session was also significant, with lower temporal summation during audio compared to silence. Temporal summation was marginally higher but not significantly different in the Music group compared to the Nature group (p=.089) indicating that participants in the Music group may have had higher temporal summation. Mechanical pain tolerance, the amount of pressure on the thumb that was rated at >80, was not significantly different between groups or between sessions (Table 3).

**Table 3.** Pain variables by session (Audio, Silence) and audio group assignment (Music, Nature)

| | Music Group<br>(*n* = 4)<br>(*M* (*SD*)) | Nature Group<br>(*n* = 5)<br>(*M* (*SD*)) | Session<br>*F* (1, 7)<br>(*p*, partial $\eta^2$) | Group<br>*F* (1, 7)<br>(*p*, partial $\eta^2$) | Group by<br>Session<br>*F* (1, 7)<br>(*p*, partial $\eta^2$) |
| --- | --- | --- | --- | --- | --- |
| Temporal Summation: Audio | 20.25 (14.29) | 4.13 (5.60) | 5.94<br>(.045*, .46) | 3.90<br>(.089<sup>†</sup>, .36) | 6.33<br>(.04*, .48) |
| Temporal Summation:<br>Silence | 20.17 (13.14) | 9.40 (7.44) |  |  |  |
| Mechanical Pain Tolerance:<br>Audio | 4.12 (1.02) | 5.17 (0.85) | 0.01<br>(.92, .002) | 2.94<br>(.13, .30) | .09<br>(.77, .01) |
| Mechanical Pain Tolerance:<br>Silence | 4.18 (1.00) | 5.05 (0.94) |  |  |  |

**Notes:** Temporal summation is the difference in pain rating out of 100 between a single stimulus and the series of ten in the non-dominant forearm. Mechanical pain tolerance is the pressure intensity (kg/cm<sup>2</sup>) at which participants rated pain in their non-dominant thumb at 70 out of 100. Pain measures were assessed with repeated measures ANOVA. Effect sizes are indicated by partial $\eta^2$ .
\*Indicates significant effects at $p < .05$ . <sup>†</sup>Indicates non-significant effects at $p < .10$ .
**Abbreviations:** PSI, pounds per square inch; M, mean; SD, standard deviation.

Adding covariates of FMness, depression, and anxiety symptoms to the ANCOVA for temporal summation (Table 4) revealed that the significant interaction of Group by Session was still present, but that there were significant interactions of Session and FM symptoms and Session and anxiety symptoms. These individual differences appear to mediate the effects of Group and Session, as the main effects for both were no longer significant.

**Table 4.** ANCOVA table for temporal summation by session (Audio, Silence) and audio group assignment (Music, Nature) corrected for FMness, depression, and anxiety symptom severity

| | Type III Sum of Squares | *df* | *F* | *p* | Partial $\eta^2$ |
| --- | --- | --- | --- | --- | --- |
| Session | 4.89 | 1 (4) | 3.39 | .14 | .46 |
| Group | 31.08 | 1 (4) | 0.13 | .74 | .03 |
| WOLFE FMness | 35.67 | 1 (4) | 0.15 | .72 | .04 |
| PROMIS Depression | 96.40 | 1 (4) | 0.40 | .56 | .09 |
| PROMIS Anxiety | 77.86 | 1 (4) | 0.32 | .60 | .07 |
| Session by Group | 9.66 | 1 (4) | 6.69 | .06<sup>†</sup> | .63 |
| Session by WOLFE FMness | 12.11 | 1 (4) | 8.39 | .04* | .68 |
| Session by PROMIS Depression | 1.87 | 1 (4) | 1.29 | .32 | .24 |
| Session by PROMIS Anxiety | 11.92 | 1 (4) | 8.25 | .045* | .67 |

**Notes:** Adjusted temporal summation measures were assessed with repeated measures ANCOVA. Effect sizes are indicated by partial $\eta^2$ . \*Indicates significant effects at $p < .05$ . <sup>†</sup>Indicates non-significant effects at $p < .10$ .
**Abbreviations:** ANCOVA, analysis of covariance; HR, heart rate; *df*, degrees of freedom; FM, fibromyalgia; PROMIS, Patient-Reported Outcomes Measurement Information System.

Adding covariates to the ANCOVA for mechanical pain tolerance (Table 5) revealed a significant interaction of Session and depression as well as non-significant trends for the main effects of Session and FM symptoms. No other effects or interactions were significant.

**Table 5.** ANCOVA table for mechanical pain tolerance by session (Audio, Silence) and audio group assignment (Music, Nature) corrected for FMness, depression, and anxiety symptom severity

| | Type III Sum of Squares | *df* | *F* | *p* | Partial $\eta^2$ |
| --- | --- | --- | --- | --- | --- |
| Session | 0.88 | 1 (4) | 4.64 | .097<sup>†</sup> | .54 |
| Group | 0.79 | 1 (4) | 1.38 | .31 | .26 |
| WOLFE FMness | 3.58 | 1 (4) | 6.29 | .07<sup>†</sup> | .61 |
| PROMIS Depression | 0.46 | 1 (4) | 0.81 | .42 | .17 |
| PROMIS Anxiety | 0.71 | 1 (4) | 1.25 | .33 | .24 |
| Session by Group | 0.34 | 1 (4) | 1.82 | .25 | .31 |
| Session by WOLFE FMness | 0.42 | 1 (4) | 2.24 | .21 | .36 |
| Session by PROMIS Depression | 2.07 | 1 (4) | 10.97 | .03* | .73 |
| Session by PROMIS Anxiety | 0.11 | 1 (4) | 0.58 | .25 | .31 |

**Notes:** Adjusted mechanical pain tolerance measures were assessed with repeated measures ANCOVA. Effect sizes are indicated by partial $\eta^2$ . \*Indicates significant effects at $p < .05$ . <sup>†</sup>Indicates non-significant effects at $p < .10$ .
**Abbreviations:** ANCOVA, analysis of covariance; HR, heart rate; *df*, degrees of freedom; FM, fibromyalgia; PROMIS, Patient-Reported Outcomes Measurement Information System.

### 3.3 ANS Measures

The ANOVA for heart rate revealed a non-significant trend for Group, with the Nature group having slightly more reduced heart rate from baseline compared to the Music group (Figure 3). No other effects were significant. The ANOVA for heart rate variability (HRV) revealed no significant effects (Figure 4, Supplementary Table 2).

**Figure 3.**
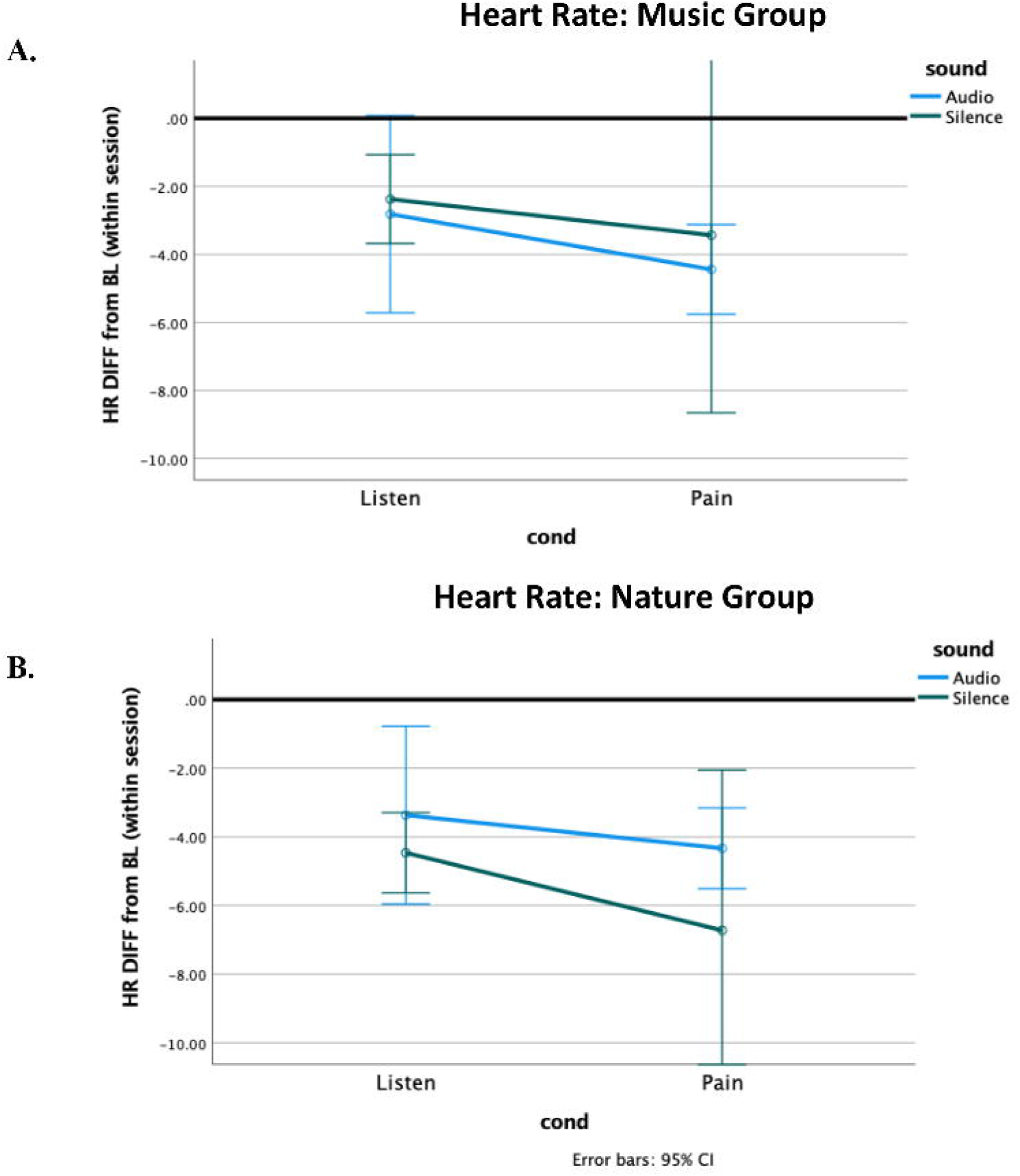
Heart Rate difference from within-session baseline. Both groups’ heart rate decreased from baseline to the Listening condition, and further decreased during Pain. The Music group had greater pain-related decrease to Music compared to silence, and the Nature group had greater pain-related decrease to Silence compared to Nature Sounds.

**Figure 4.**
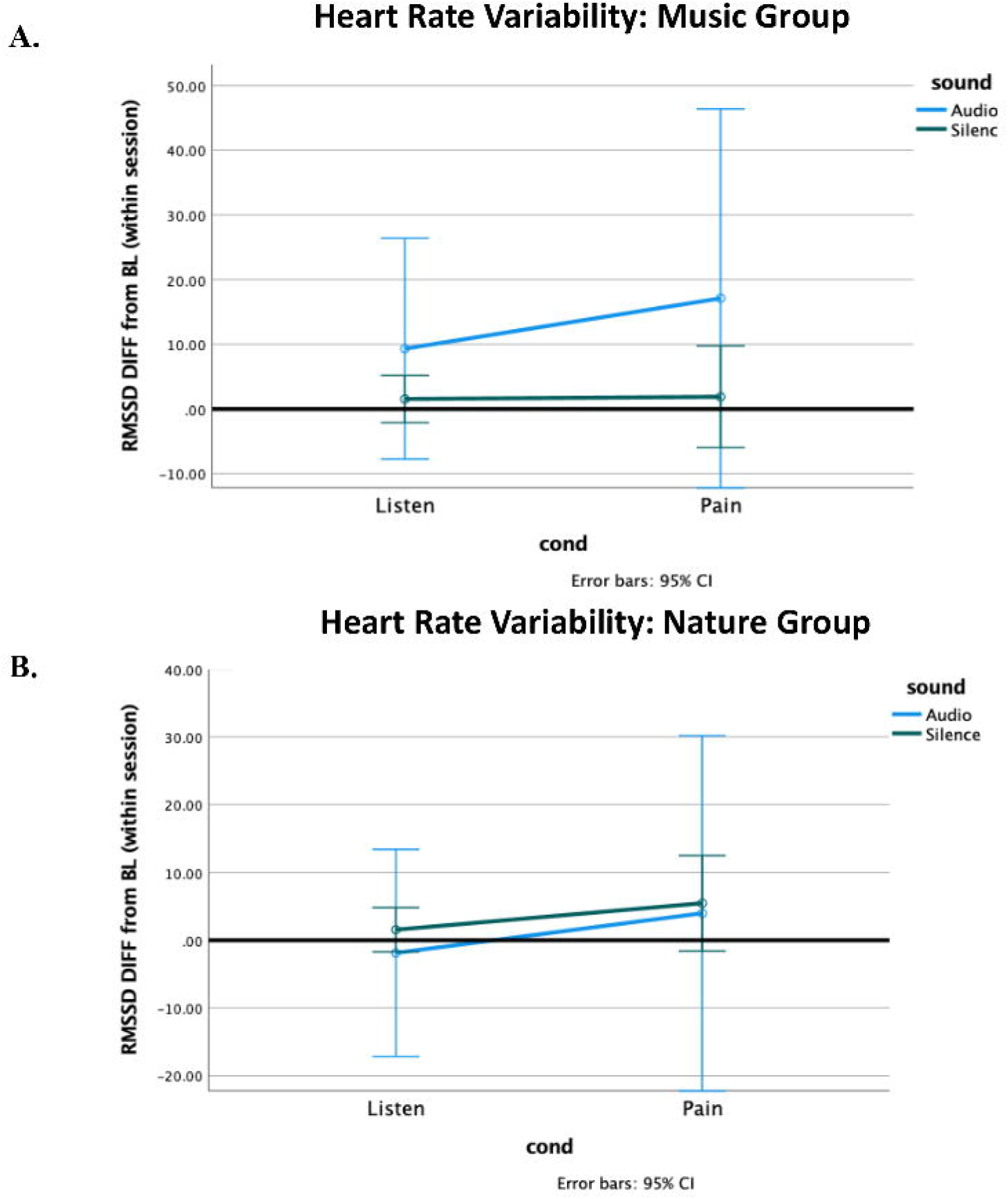
Heart Rate Variability (HRV) difference from within-session baseline. The Music group’s HRV increased from baseline to the Listening condition, and further increased during Pain, with no effect observed for Silence. The Nature group had marginally greater HRV during Pain. The HRV during Pain was associated with a high standard deviation in both groups.

However, the addition of FMness, depression, and anxiety symptoms in the ANCOVA for heart rate (Supplementary Table 3) yielded several significant effects and interactions. The main effects of Condition (Listen versus Pain) and Session (Audio versus Silence) were both significant, but the main effect for Group was no longer near the significance threshold. The two-way interactions of Condition by anxiety and Session by anxiety were significant, as was the Session by FM symptoms. The interaction of Condition by FM symptoms was marginal. Finally, the three-way interaction of Session by Condition by anxiety was significant and Session by Condition by depression was marginal though not significant, suggesting that mood symptoms were impacting how heart rate changed with pain compared to listening alone. No significant effects or interactions were found in the ANCOVA for HRV (Supplementary Table 4).

### 3.4 Correlations

#### 3.4.1 Fibromyalgia, Mood, and Music Experience

Correlations between FMness, depression, anxiety symptoms, music experience scores and between-session changes in the primary outcomes of temporal pain summation, mechanical pain tolerance, heart rate and heart rate variability are presented in Supplementary Table 5. FM questionnaires were related to one another, but not with the pain or ANS outcome measures. FMness was negatively correlated with both depression and anxiety symptoms, though neither reached significance. FM symptoms were also negatively correlated with affective reactions to music, and FMness was positively correlated with commitment to music. Depression was negatively correlated with commitment to music and innovative musical aptitude. Anxiety was strongly positively correlated with affective reactions to music. Innovative musical aptitude and positive psychotropic effects were strongly positively correlated with reactive musical behavior.

#### 3.4.2 Pain and ANS Response with Mood and Music Experience

Anxiety was related to both temporal pain summation and heart rate, such that greater anxiety was associated with a smaller change in temporal summation and a smaller change in heart rate during pain. Temporal pain summation change between sessions was positively correlated with reactive musical behavior. There was a marginal negative correlation between the change in mechanical pain tolerance and depression, such that the higher the depression symptoms, the less tolerance changed between audio and silence. There was also a significant positive correlation between positive psychotropic effects from music and the change in mechanical pain tolerance, such that those who had greater psychotropic effects had a greater change in mechanical pain tolerance between audio and silence. Heart rate change with pain between sessions was negatively correlated with affective reactions to music. HRB change with pain between sessions was negatively correlated with positive psychotropic effects from music and positively correlated with between session change in temporal pain summation, such that greater change in heart rate variability was associated with greater change in temporal summation between sessions.

## 4 Discussion

In this pilot study, we measured the analgesic effects associated with music and nature sounds on objective autonomic system responsiveness to painful stimuli. Our experimental design allowed for blinding during both data collection and analysis, reducing the potential for bias. By counterbalancing the order of audio presentation, we showed feasibility of repeated measures testing in patients with FM while controlling for order effects. Even in our small sample, randomization successfully yielded relatively matched groups, with no group differences observed for FM symptoms, age, marital status, or education. By random chance, we did observe between group differences on anxiety, although the Nature group was numerically only 7 points higher than the Music group, and both groups were within one standard deviation of the standardized mean on the PROMIS scale.

### 4.1 Objective Pain

This study aimed to manipulate two potential mechanisms for music-evoked analgesia: cognitive distraction and physiological or vagal response alteration. By using musical stimuli and an active control (Nature sounds), and comparing to each participant’s own Silence control, our experimental design allows for examination of distraction due to general relaxing audio, as well as examining music-specific analgesia by comparing the Music to the Nature Sound condition directly. We also examined the relationship to individual differences on FM severity, anxiety and depression symptoms, and variability in music experience. We hypothesized that both listening conditions (Music and Nature) would reduce pain sensitivity compared to testing during Silence and that Music listening would reduce pain sensitivity compared to Nature sounds.(30-32)

#### 4.1.1 Temporal Summation

We observed a strong effect of cognitive distraction, with reduced temporal summation during either audio condition compared to silence, indicating that the auditory stimulus was effective in reducing pain. The direction of the group difference was opposite to our hypothesis, with the Nature group showing an analgesic effect, while the Music group showed none. This could be due to the confound of anxiety symptoms between the groups, or it could be a potential confound of pre-existing differences in sensitization between the groups, as temporal summation overall was somewhat higher in the Music group compared to the Nature group.(23)

#### 4.1.2 Mechanical Pain Tolerance

Interestingly, we did not observe any effects of group or condition on tolerance to thumb pressure. This was surprising as this test usually shows high sensitivity for variations in pain response.(25) However, it is possible that the transient changes between sessions were too small to be observed in this small sample, and that a larger sample or longer intervention would be necessary to see differences in maximal pain tolerance.

### 4.2 ANS

#### 4.2.1 Heart Rate

We also hypothesized that Music would increase vagal input to the autonomic nervous system, decreasing heart rate and increasing heart rate variability compared to both Silence and Nature sounds. Vagal response during pain is a coping mechanism (57). We did observe a difference between the groups on heart rate pointing to the feasibility of the chosen stimuli, yet the direction was opposite to our hypothesis with the Nature group having greater reductions from baseline compared to the Music group. This could also be related to group differences in anxiety or other pre-existing physiological differences between the groups. The Nature group, having higher anxiety (*p* = 0.03), could have had elevated heart rate at baseline, thereby having more chance for the analgesic effect to be observed. In our analysis, we corrected for the within-session baseline to address this possibility.

#### 4.2.2 Heart Rate Variability

HRV is a better measure for vagal response than raw heart rate.(58) However, we observed no significant effects for HRV, suggesting that we were underpowered to observe a vagal response with this small sample. While not significant, the Music group did show a pattern of response that was consistent with vagal activation similar to other studies, with reduction from baseline and then further reduction during pain, that was not observed in the Nature group.(59, 60) Such anticipated response might be due to emotional expression towards the music stimulus, enjoyment, or simply just being entertained. A larger sample would be needed to clarify whether there is a greater vagal response to music.

### 4.3 Individual Differences

Individual differences likely play a role in how a person will respond to auditory stimulation.(61-63) In this study, we hypothesized that analgesic responsiveness would vary dependent on symptoms of FMness, anxiety, and depression.(54) We therefore examined correlations between the outcomes of interest and various individual difference measures to elucidate the potential relationships between them.

#### 4.3.1 FM symptoms

In our sample of participants, we did not find relationships with FM symptoms and objective pain or ANS measures, contrary to what we had hypothesized. However, we did see weak negative correlations with an aggregate metric of FM symptoms, FMness, and both depression and anxiety mood symptoms. In our sample, those with higher FMness reported lower depression and anxiety symptoms. Although not significant, these relationships underscore the fuzzy boundaries and complex relationships between FMness and mood symptoms. Depression specifically can be associated with both increased and decreased physical motor symptoms.(64) These relationships should be explored further to determine whether a specific profile of mood symptoms is related to analgesic responsiveness in people with FM. Moreover, and on a broader scale, physical and emotional symptoms should also be explored to determine their relationship with analgesic responsiveness.

#### 4.3.2 Music Experience

The Music Experience Questionnaire (MEQ) revealed several interesting relationships. We observed that people with greater FM symptoms reported weaker affective reactions to music, or alternatively, that the less people reported responding to music affectively, the greater their FM symptoms. However, the aggregate FMness measure was positively related to commitment to music, suggesting that even though people with more severe FM symptoms may be having weaker affective responses to music, they still report a deep commitment and importance of music in their lives. This may in part underlie the positive effects of music interventions for FM.(32, 42, 65)

Mood symptoms have previously been related to the MEQ.(55) In our sample, we saw a negative relationship between depression and commitment to music and innovative musical aptitude. This is consistent with symptoms of depression of anhedonia and self-criticism. As the innovative musical aptitude is self-reported and not objectively measured, it is impossible to know how accurate that assessment was. The relationship with depression suggests that participants may have been more critical of their own abilities in this area compared to those who are not experiencing depressive symptoms.

A positive relationship was observed between anxiety and affective reactions to music. As anxiety is often associated with labile emotional reactions, strong affective responses to music would be consistent with that. This relationship may also be underlying some of our other findings associated with anxiety. More research is necessary to determine whether people with higher anxiety would benefit more from an auditory listening analgesia intervention. (66-68)

Finally, there were some relationships between measures of the MEQ showing that innovative musical aptitude and positive psychotropic effects from music were both strongly positively correlated with reactive musical behavior. These relationships are perhaps unsurprising from a reward perspective where those who get intrinsic reward from music are more likely to engage in it.(69) However, this preliminary study could not suggest who might benefit from music more than nature sounds. Future studies are needed to determine whether specific patient characteristics would allow some to benefit more from music compared to nature sounds.

#### 4.3.3 Anxiety, Depression

We observed significant effects of anxiety on temporal summation. In the ANCOVA analysis, participants with higher anxiety had greater analgesic effects during the audio listening session. In our correlation analysis, we observed that higher anxiety was associated with smaller changes in temporal summation and smaller changes in heart rate during pain, which indicates those with higher anxiety had reduced analgesic effect of distraction. This is potentially an important finding, as anxiety is a common clinical presentation for people with fibromyalgia and chronic pain and may influence treatment choices.(70, 71) In our sample, anxiety was randomly different between our two audio groups, with those in the Nature group having higher anxiety. Unfortunately, this confound means that we cannot completely separate the effects of anxiety from the response to Music versus Nature sounds and will be important to address in future studies.

We also observed a significant relationship between depression and analgesic effects for mechanical pain tolerance, with depression blunting the analgesic effect from the audio condition. Depression often includes physical manifestations, including blunting of energy and increased tiredness. The relationship with mechanical pain tolerance could be a sign of those physical manifestations. In our correlation analysis, higher depression symptoms were related to less change in mechanical pain tolerance between sessions, which could be related to sensory symptoms of depression. However, the PROMIS depression measures that we used in this study are not sensitive enough to capture depression subtypes.

Perhaps unsurprisingly, mood symptoms significantly impacted heart rate. When the effects of anxiety and depression were controlled, significant effects of pain and audio session were revealed that were not observed in the uncorrected ANOVA models. However, the effect of Group was no longer significant, indicating that there were no additional analgesic effects of music over nature sounds, although, as for temporal summation, these results are limited by the confounding of anxiety with random group assignment. Further studies are needed to clarify these relationships.

Finally, we observed that those with greater behavioral reactions to music, as measured by the MEQ, had greater change in temporal summation scores between sessions, regardless of whether they were listening to music or nature sounds, and those who reported experiencing greater psychotropic effects from music had greater changes in mechanical pain tolerance between sessions.

## 5 Limitations

This study is limited by the small sample size, and results should be interpreted with caution. Additionally, our groups differed on anxiety potentially confounding our results, although all participants were in the mild-moderate anxiety range. However, inherently when having a small sample size, it is somewhat easier to detect within-participant effects rather than between-participant effects. The effects we observed relating to anxiety, depression, and FMness symptoms indicate the need for future studies to include these factors in the study design. Our experimental design using nature sounds as an auditory control and carefully selected musical selections with characteristics hypothesized to facilitate relaxation and analgesia is a strength, and one that can be used in future studies to separate the effects of auditory distraction from music-specific effects.

## 6 Conclusion

In conclusion, our current results did not support our hypothesis of stronger analgesic effects of music versus distracting nature sounds, however, we did observe strong effects of auditory distraction on pain temporal summation and tolerance. The confounding effect of anxiety symptoms in our study, as well as the individual differences observed on the MEQ, suggest that variability in mood and other factors may be important in understanding how individuals will respond to music or other auditory stimuli to gain therapeutic analgesic effects. While these results should be treated with caution, this study provides preliminary evidence that some individuals may benefit from music or audio stimulation as a treatment more than others. Further study is warranted.

## Data Availability

All data produced are available online at https://osf.io/84baz.

## 8 Author Contributions

**Rebecca J. Lepping**: Conceptualization, Methodology, Formal analysis, Investigation, Data Curation, Writing - Original Draft, Project administration, Funding acquisition, **Miranda L. McMillan**: Formal analysis, Investigation, Data Curation, Writing - Review & Editing, Project administration, **Andrea L. Chadwick**: Conceptualization, Methodology, Resources, Writing - Original Draft, Writing - Review & Editing, Funding acquisition, **Zaid M. Mansour**: Writing - Review & Editing, **Laura E. Martin**: Resources, Writing - Review & Editing, **Kathleen M. Gustafson**: Conceptualization, Methodology, Investigation, Resources, Writing - Review & Editing

## 10 Acknowledgments

We humbly thank the participants for contributing to the study.

## 11 Data Availability Statement

This trial and protocol are registered at ClinicalTrials.gov NCT04059042 and Open Science Framework (OSF) Preregistration https://osf.io/84baz. The datasets generated and analyzed for this study can be found in the OSF website https://osf.io/84baz.

## References

1. Williams DA, Clauw DJ. Understanding fibromyalgia: lessons from the broader pain research community. The journal of pain : official journal of the American Pain Society. 2009;10(8):777–91.

2. Woolf CJ. Central sensitization: implications for the diagnosis and treatment of pain. Pain. 2011;152(3 Suppl):S2–15.

3. Melemedjian OK, Tillu DV, Asiedu MN, Mandell EK, Moy JK, Blute VM, et al. BDNF regulates atypical PKC at spinal synapses to initiate and maintain a centralized chronic pain state. Molecular pain. 2013;9:12.

4. Clauw DJ. Fibromyalgia: A Clinical Review. JAMA. 2014;311(5):1547–55.

5. Aasvang EK, Gmaehle E, Hansen JB, Gmaehle B, Forman JL, Schwarz J, et al. Predictive risk factors for persistent postherniotomy pain. Anesthesiology. 2010;112(4):957–69.

6. Arendt-Nielsen L, Yarnitsky D. Experimental and clinical applications of quantitative sensory testing applied to skin, muscles and viscera. The journal of pain : official journal of the American Pain Society. 2009;10(6):556–72.

7. Yarnitsky D, Crispel Y, Eisenberg E, Granovsky Y, Ben-Nun A, Sprecher E, et al. Prediction of chronic post-operative pain: pre-operative DNIC testing identifies patients at risk. Pain. 2008;138(1):22–8.

8. Ablin K, Clauw DJ. From fibrositis to functional somatic syndromes to a bell-shaped curve of pain and sensory sensitivity: evolution of a clinical construct. Rheum Dis Clin North Am. 2009;35(2):233–51.

9. Diatchenko L, Nackley AG, Slade GD, Bhalang K, Belfer I, Max MB, et al. Catechol-O-methyltransferase gene polymorphisms are associated with multiple pain-evoking stimuli. Pain. 2006;125(3):216–24.

10. Gwilym SE, Keltner JR, Warnaby CE, Carr AJ, Chizh B, Chessell I, et al. Psychophysical and functional imaging evidence supporting the presence of central sensitization in a cohort of osteoarthritis patients. Arthritis and rheumatism. 2009;61(9):1226–34.

11. Nielsen CS, Staud R, Price DD. Individual differences in pain sensitivity: measurement, causation, and consequences. The journal of pain : official journal of the American Pain Society. 2009;10(3):231–7.

12. Neddermeyer TJ, Fluhr K, Lotsch J. Principle components analysis of pain thresholds to thermal, electrical, and mechanical stimuli suggests a predominant common source of variance. Pain. 2008;138(2):286–91.

13. Fillingim RB. Individual differences in pain responses. Curr Rheumatol Rep. 2005;7(5):342–7.

14. Kleinbohl D, Holzl R, Moltner A, Rommel C, Weber C, Osswald PM. Psychophysical measures of sensitization to tonic heat discriminate chronic pain patients. Pain. 1999;81(1-2):35–43.

15. Wilder-Smith OHG, Tassonyi E, Arendt-Nielsen L. Preoperative back pain is associated with diverse manifestations of central neuroplasticity. Pain. 2002;97(3):189–94.

16. Whitehead WE, Holtkotter B, Enck P, Hoelzl R, Holmes KD, Anthony J, et al. Tolerance for rectosigmoid distention in irritable bowel syndrome. Gastroenterology. 1990;98(5 Pt 1):1187–92.

17. Gibson SJ, Littlejohn GO, Gorman MM, Helme RD, Granges G. Altered heat pain thresholds and cerebral event-related potentials following painful CO2 laser stimulation in subjects with fibromyalgia syndrome. Pain. 1994;58(2):185–93.

18. Giesecke J, Reed BD, Haefner HK, Giesecke T, Clauw DJ, Gracely RH. Quantitative sensory testing in vulvodynia patients and increased peripheral pressure pain sensitivity. Obstetrics and gynecology. 2004;104(1):126–33.

19. Gracely RH, Petzke F, Wolf JM, Clauw DJ. Functional magnetic resonance imaging evidence of augmented pain processing in fibromyalgia. Arthritis and rheumatism. 2002;46(5):1333–43.

20. Greenspan JD, Slade GD, Bair E, Dubner R, Fillingim RB, Ohrbach R, et al. Pain sensitivity risk factors for chronic TMD: descriptive data and empirically identified domains from the OPPERA case control study. The journal of pain : official journal of the American Pain Society. 2011;12(11 Suppl):T61–74.

21. Leffler AS, Hansson P, Kosek E. Somatosensory perception in a remote pain-free area and function of diffuse noxious inhibitory controls (DNIC) in patients suffering from long-term trapezius myalgia. Eur J Pain. 2002;6(2):149–59.

22. Clauw DJ, Chrousos GP. Chronic pain and fatigue syndromes: overlapping clinical and neuroendocrine features and potential pathogenic mechanisms. Neuroimmunomodulation. 1997;4(3):134–53.

23. Hsu MC, Harris RE, Sundgren PC, Welsh RC, Fernandes CR, Clauw DJ, et al. No consistent difference in gray matter volume between individuals with fibromyalgia and age-matched healthy subjects when controlling for affective disorder. Pain. 2009;143(3):262–7.

24. As-Sanie S, Harris RE, Harte SE, Tu FF, Neshewat G, Clauw DJ. Increased pressure pain sensitivity in women with chronic pelvic pain. Obstetrics and gynecology. 2013;122(5):1047–55.

25. Petzke F, Harris RE, Williams DA, Clauw DJ, Gracely RH. Differences in unpleasantness induced by experimental pressure pain between patients with fibromyalgia and healthy controls. Eur J Pain. 2005;9(3):325–35.

26. Ribeiro MKA, Alcantara-Silva TRM, Oliveira JCM, Paula TC, Dutra JBR, Pedrino GR, et al. Music therapy intervention in cardiac autonomic modulation, anxiety, and depression in mothers of preterms: randomized controlled trial. BMC Psychol. 2018;6(1):57.

27. Witvliet CVO, Vrana SR. Play it again Sam: Repeated exposure to emotionally evocative music polarises liking and smiling responses, and influences other affective reports, facial EMG, and heart rate. Cognition & Emotion. 2007;21(1):3–25.

28. Williams C, Hine T. An investigation into the use of recorded music as a surgical intervention: A systematic, critical review of methodologies used in recent adult controlled trials. Complement Ther Med. 2018;37:110–26.

29. Hole J, Hirsch M, Ball E, Meads C. Music as an aid for postoperative recovery in adults: a systematic review and meta-analysis. Lancet. 2015;386(10004):1659–71.

30. Fleming PS, Strydom H, Katsaros C, MacDonald L, Curatolo M, Fudalej P, et al. Non-pharmacological interventions for alleviating pain during orthodontic treatment. Cochrane Database Syst Rev. 2016;12:CD010263.

31. Basinski K, Zdun-Ryzewska A, Majkowicz M. The Role of Musical Attributes in Music-Induced Analgesia: A Preliminary Brief Report. Front Psychol. 2018;9:1761.

32. Garza-Villarreal EA, Pando V, Vuust P, Parsons C. Music-Induced Analgesia in Chronic Pain Conditions: A Systematic Review and Meta-Analysis. Pain Physician. 2017;20(7):597–610.

33. Lunde SJ, Vuust P, Garza-Villarreal EA, Vase L. Music-induced analgesia: how does music relieve pain? Pain. 2019;160(5):989–93.

34. Villarreal EA, Brattico E, Vase L, Ostergaard L, Vuust P. Superior analgesic effect of an active distraction versus pleasant unfamiliar sounds and music: the influence of emotion and cognitive style. Plos One. 2012;7(1):e29397.

35. Juslin PN, Sloboda JA. Music and emotion : theory and research. Oxford ; New York: Oxford University Press; 2001. viii, 487 p. p.

36. Reybrouck M, Eerola T. Music and Its Inductive Power: A Psychobiological and Evolutionary Approach to Musical Emotions. Front Psychol. 2017;8:494.

37. Aselton P. Sources of stress and coping in American college students who have been diagnosed with depression. J Child Adolesc Psychiatr Nurs. 2012;25(3):119–23.

38. Brandes V, Terris DD, Fischer C, Loerbroks A, Jarczok MN, Ottowitz G, et al. Receptive music therapy for the treatment of depression: a proof-of-concept study and prospective controlled clinical trial of efficacy. Psychother Psychosom. 2010;79(5):321–2.

39. Blood AJ, Zatorre RJ. Intensely pleasurable responses to music correlate with activity in brain regions implicated in reward and emotion. Proc Natl Acad Sci U S A. 2001;98(20):11818–23.

40. Salimpoor VN, Benovoy M, Larcher K, Dagher A, Zatorre RJ. Anatomically distinct dopamine release during anticipation and experience of peak emotion to music. Nat Neurosci. 2011;14(2):257–62.

41. Martin-Saavedra JS, Vergara-Mendez LD, Pradilla I, Velez-van-Meerbeke A, Talero-Gutierrez C. Standardizing music characteristics for the management of pain: A systematic review and meta-analysis of clinical trials. Complement Ther Med. 2018;41:81–9.

42. Garza-Villarreal EA, Wilson AD, Vase L, Brattico E, Barrios FA, Jensen TS, et al. Music reduces pain and increases functional mobility in fibromyalgia. Front Psychol. 2014;5:90.

43. Garza-Villarreal EA, Jiang Z, Vuust P, Alcauter S, Vase L, Pasaye EH, et al. Music reduces pain and increases resting state fMRI BOLD signal amplitude in the left angular gyrus in fibromyalgia patients. Front Psychol. 2015;6:1051.

44. Harris PA, Taylor R, Thielke R, Payne J, Gonzalez N, Conde JG. Research electronic data capture (REDCap)--a metadata-driven methodology and workflow process for providing translational research informatics support. J Biomed Inform. 2009;42(2):377–81.

45. Harris PA, Taylor R, Minor BL, Elliott V, Fernandez M, O’Neal L, et al. The REDCap consortium: Building an international community of software platform partners. J Biomed Inform. 2019;95:103208.

46. Brummett CM, Janda AM, Schueller CM, Tsodikov A, Morris M, Williams DA, et al. Survey Criteria for Fibromyalgia Independently Predict Increased Postoperative Opioid Consumption after Lower-extremity Joint Arthroplasty: A Prospective, Observational Cohort Study. Anesthesiology. 2013;119(6):1434–43.

47. Janda AM, As-Sanie S, Rajala B, Tsodikov A, Moser SE, Clauw DJ, et al. Fibromyalgia survey criteria are associated with increased postoperative opioid consumption in women undergoing hysterectomy. Anesthesiology. 2015;122(5):1103–11.

48. Wolfe F, Clauw DJ, Fitzcharles MA, Goldenberg DL, Hauser W, Katz RS, et al. Fibromyalgia criteria and severity scales for clinical and epidemiological studies: a modification of the ACR Preliminary Diagnostic Criteria for Fibromyalgia. The Journal of rheumatology. 2011;38(6):1113–22.

49. Wolfe F. Fibromyalgianess. Arthritis and rheumatism. 2009;61(6):715–6.

50. Tan G, Jensen MP, Thornby JI, Shanti BF. Validation of the Brief Pain Inventory for chronic nonmalignant pain. The journal of pain : official journal of the American Pain Society. 2004;5(2):133–7.

51. Jensen MP, Turner JA, Romano JM, Fisher LD. Comparative reliability and validity of chronic pain intensity measures. Pain. 1999;83(2):157–62.

52. Dworkin RH, Turk DC, Wyrwich KW, Beaton D, Cleeland CS, Farrar JT, et al. Interpreting the clinical importance of treatment outcomes in chronic pain clinical trials: IMMPACT recommendations. The journal of pain : official journal of the American Pain Society. 2008;9(2):105–21.

53. Bennett RM, Friend R, Jones KD, Ward R, Han BK, Ross RL. The Revised Fibromyalgia Impact Questionnaire (FIQR): validation and psychometric properties. Arthritis research & therapy. 2009;11(4):R120.

54. Giesecke T, Williams DA, Harris RE, Cupps TR, Tian X, Tian TX, et al. Subgrouping of fibromyalgia patients on the basis of pressure-pain thresholds and psychological factors. Arthritis and rheumatism. 2003;48(10):2916–22.

55. Werner PD, Swope AJ, Heide FJ. The Music Experience Questionnaire: development and correlates. J Psychol. 2006;140(4):329–45.

56. Wolfe F, Smythe HA, Yunus MB, Bennett RM, Bombardier C, Goldenberg DL, et al. The American College of Rheumatology 1990 Criteria for the Classification of Fibromyalgia. Report of the Multicenter Criteria Committee. Arthritis and rheumatism. 1990;33(2):160–72.

57. Fabes RA, Eisenberg N. Regulatory control and adults’ stress-related responses to daily life events. J Pers Soc Psychol. 1997;73(5):1107–17.

58. Nussinovitch U, Elishkevitz KP, Katz K, Nussinovitch M, Segev S, Volovitz B, et al. Reliability of Ultra-Short ECG Indices for Heart Rate Variability. Ann Noninvasive Electrocardiol. 2011;16(2):117–22.

59. Forte G, Troisi G, Pazzaglia M, Pascalis V, Casagrande M. Heart Rate Variability and Pain: A Systematic Review. Brain Sci. 2022;12(2).

60. Koelsch S, Jancke L. Music and the heart. Eur Heart J. 2015;36(44):3043–9.

61. Martarelli CS, Mayer B, Mast FW. Daydreams and trait affect: The role of the listener’s state of mind in the emotional response to music. Conscious Cogn. 2016;46:27–35.

62. Carlson E, Burger B, London J, Thompson MR, Toiviainen P. Conscientiousness and Extraversion relate to responsiveness to tempo in dance. Hum Mov Sci. 2016;49:315–25.

63. Eerola T, Vuoskoski JK, Kautiainen H. Being Moved by Unfamiliar Sad Music Is Associated with High Empathy. Front Psychol. 2016;7:1176.

64. First MB, Spitzer, Robert L, Gibbon Miriam, and Williams, Janet B.W. Structured Clinical Interview for DSM-IV-TR Axis I Disorders, Research Version, Non-patient Edition. (SCID-I/NP). New York: Biometrics Research, New York State Psychiatric Institute; 2002.

65. Onieva-Zafra MD, Castro-Sanchez AM, Mataran-Penarrocha GA, Moreno-Lorenzo C. Effect of music as nursing intervention for people diagnosed with fibromyalgia. Pain Manag Nurs. 2013;14(2):e39–46.

66. Vuong V, Mosabbir A, Paneduro D, Picard L, Faghfoury H, Evans M, et al. Effects of Rhythmic Sensory Stimulation on Ehlers-Danlos Syndrome: A Pilot Study. Pain Res Manag. 2020;2020:3586767.

67. Torres E, Pedersen IN, Perez-Fernandez JI. Randomized Trial of a Group Music and Imagery Method (GrpMI) for Women with Fibromyalgia. J Music Ther. 2018;55(2):186–220.

68. Guetin S, Ginies P, Siou DK, Picot MC, Pommie C, Guldner E, et al. The effects of music intervention in the management of chronic pain: a single-blind, randomized, controlled trial. Clin J Pain. 2012;28(4):329–37.

69. Varella MAC. Evolved Features of Artistic Motivation: Analyzing a Brazilian Database Spanning Three Decades. Front Psychol. 2021;12:769915.

70. Aaron LA, Burke MM, Buchwald D. Overlapping conditions among patients with chronic fatigue syndrome, fibromyalgia, and temporomandibular disorder. Arch Intern Med. 2000;160(2):221–7.

71. Yunus MB. Fibromyalgia and overlapping disorders: the unifying concept of central sensitivity syndromes. Semin Arthritis Rheum. 2007;36(6):339–56.

