## Supplementary Tables for "Autonomic Nervous System Markers of Music-Elicited Analgesia in People with Fibromyalgia: A Double-Blind Randomized Pilot Study"

Supplementary Material

**Supplementary Table 1** Audio recordings used during the listening conditions.

| Year | Title | Composer | Recording Artist | Album |
| --- | --- | --- | --- | --- |
| Nature Sounds | | | | |
| 2011 | Gentle Birds and Forest Stream for Relaxation Meditation | N/A | N/A | Bird Sounds – Morning Birds for Relaxation, Meditation, Yoga, Naturescapes, Forest Ambience and Spa |
| 2011 | A Forest Ambience – Nocturne Nature Sounds Mother Earth Sleep Music for Relaxation, Meditation, Massage, Toga Class, Tai Chi, Reiki, Tantra and Zen | N/A | N/A | The Secret Garden Nature Sounds – Relaxing Sounds of Nature for Deep Sleep, Baby Sleep, Yoga Pregnancy, Yoga Music Relaxation |
| Western Classical Music | | | | |
| 2017 | Deux arabesques, L. 66: No. 1 in E Major, Andantino con moto | Debussy, Claude | Battel, Giovanni Umberto | Classical Music Collection |
| 2019 | Bagatelle No. 25 in A Minor, WoO 59 “Für Elise” | Beethoven, Ludwig van | Battel, Giovanni Umberto | 50 Most Famous Pieces of Classical Music |
| 2019 | The Four Seasons, Violin Concerto No. 4 in F Minor, RV 297 “Winter”: II. Largo | Vivaldi, Antonio | Pavel Lyubomudrov, Metamorphose String Orchestra, Yuliya Lebedenko Conductor | Baroque Music for Studying & Brain Power |
| 2017 | Piano Sonata No. 14 in C-Sharp Minor, Op. 27, No. 2 “Moonlight Sonata”: I. Adagio sostenuto | Beethoven, Ludwig van | Battel, Giovanni Umberto | Sad, Melancholic Classical Music |
| 2017 | Symphony No. 4 in A Major, Op. 90 “Italian”: II. Andante con moto | Mendelssohn, Felix | Orchestra da Camera Florentina, Giuseppe Lanzetta Conductor | Romantic Music – Classical Music from the Romantic Period |

**Supplementary Table 2** Heart rate variables corrected for within session baseline by condition (Listen, Pain), session (Audio, Silence), and audio group assignment (Music, Nature).

|  | Music Group (*n* = 4)  (*M* (*SD*)) | Nature Group (*n* = 5)  (*M* (*SD*)) | Condition  *F* (1, 7)  (*p*, partial η^2^) | Session  *F* (1, 7)  (*p*, partial η^2^) | Group  *F* (1, 7)  *(p*, partial η^2^) | Group by Condition  *F* (1, 7) *(p*, partial η^2^) | Group by Session  *F* (1, 7) *(p*, partial η^2^) | Condition by Session  *F* (1, 7) *(p*, partial η^2^) | 3-way interaction  *F* (1, 7) *(p*, partial η^2^) |
| --- | --- | --- | --- | --- | --- | --- | --- | --- | --- |
| HR_BL_: Cond. = Listen  Session = Audio | -2.81 (1.78) | -3.37 (2.85) | 1.65  (.24, .19) | 0.25  (.63, .03) | 3.76  (.09^†^, .35) | 0.01  (.91, .002) | 1.46  (.27, .17) | 0.16  (.71, .02) | 1.02  (.35, .13) |
| HR_BL_: Cond. = Pain  Session = Audio | -4.44 (1.30) | -4.33 (0.95) |  |  |  |  |  |  |  |
| HR_BL_: Cond. = Listen  Session = Silence | -2.37 (0.69) | -4.46 (1.33) |  |  |  |  |  |  |  |
| HR_BL_: Cond. = Pain  Session = Silence | -3.43 (1.92) | -6.72 (5.60) |  |  |  |  |  |  |  |
| HRV_BL_: Cond. = Listen  Session = Audio | 9.33 (21.46) | -1.87 (4.41) | 2.92  (.13, .29) | 0.37  (.57, .05) | 0.87  (.38, .11) | 0.02  (.88, .003) | 0.87  (.38, .11) | 1.18  (.31, .14) | 0.40  (.55, .06) |
| HRV_BL_: Cond. = Pain  Session = Audio | 17.11 (37.67) | 3.99 (3.19) |  |  |  |  |  |  |  |
| HRV_BL_: Cond. = Listen  Session = Silence | 1.55 (3.71) | 1.56 (2.52) |  |  |  |  |  |  |  |
| HRV_BL_: Cond. = Pain  Session = Silence | 1.89 (6.73) | 5.46 (6.60) |  |  |  |  |  |  |  |

**Notes:** Heart rate measures were assessed with repeated measures ANOVA. ^†^Indicates non-significant effects at *p*<.10.

**Abbreviations:** M, mean; SD, standard deviation; HR_BL_, heart rate baseline corrected; Cond., condition; HRV, heart rate variability: root mean square of successive differences between heartbeats baseline corrected.

**Supplementary Table 3** ANCOVA table for heart rate (HR_BL_) corrected for within session baseline by condition (Listen, Pain), session (Audio, Silence), and audio group assignment (Music, Nature) corrected for fibromyalgia, depression, and anxiety symptom severity.

|  | Type III Sum of Squares | *df* | *F* | *p* | Partial η^2^ |
| --- | --- | --- | --- | --- | --- |
| Condition | 33.70 | 1 (4) | 7.06 | .057^†^ | .64 |
| Session | 37.99 | 1 (4) | 25.87 | .007* | .87 |
| Group | 4.30 | 1 (4) | 0.86 | .41 | .18 |
| WOLFE FMness | 0.55 | 1 (4) | 0.11 | .76 | .03 |
| PROMIS Depression | 4.84 | 1 (4) | 0.97 | .38 | .20 |
| PROMIS Anxiety | 5.85 | 1 (4) | 1.17 | .34 | .23 |
| Condition by Session | 0.78 | 1 (4) | 1.59 | .28 | .28 |
| Condition by Group | 11.89 | 1 (4) | 2.49 | .19 | .38 |
| Condition by WOLFE FMness | 23.31 | 1 (4) | 4.89 | .09^†^ | .55 |
| Condition by PROMIS Depression | 0.03 | 1 (4) | 0.01 | .94 | .002 |
| Condition by PROMIS Anxiety | 55.38 | 1 (4) | 11.61 | .03* | .74 |
| Session by Group | 4.84 | 1 (4) | 3.29 | .14 | .45 |
| Session by WOLFE FMness | 14.24 | 1 (4) | 9.70 | .04* | .71 |
| Session by PROMIS Depression | 0.99 | 1 (4) | 0.67 | .46 | .14 |
| Session by PROMIS Anxiety | 58.18 | 1 (4) | 39.63 | .003* | .91 |
| Session by Condition by Group | 0.33 | 1 (4) | 0.66 | .46 | .14 |
| Session by Condition by WOLFE FMness | 0.08 | 1 (4) | 0.16 | .71 | .04 |
| Session by Condition by PROMIS Depression | 2.39 | 1 (4) | 4.85 | .09^†^ | .55 |
| Session by Condition by PROMIS Anxiety | 5.68 | 1 (4) | 11.53 | .03* | .74 |

**Notes:** Adjusted heart rate measures were assessed with repeated measures ANCOVA. *Indicates significant effects at *p*<.05. ^†^Indicates non-significant effects at *p*<.10.

**Abbreviations:** ANCOVA, analysis of covariance; HR_BL_, heart rate baseline corrected; df, degrees of freedom; FM, fibromyalgia; PROMIS, Patient-Reported Outcomes Measurement Information System.
**Supplementary Table 4** ANCOVA table for heart rate variability (HRV_BL_) corrected for within session baseline by condition (Listen, Pain), session (Audio, Silence), and audio group assignment (Music, Nature) corrected for fibromyalgia, depression, and anxiety symptom severity.

|  | Type III Sum of Squares | *df* | *F* | *p* | Partial η^2^ |
| --- | --- | --- | --- | --- | --- |
| Condition | 60.67 | 1 (4) | 0.71 | .45 | .15 |
| Session | 337.33 | 1 (4) | 0.59 | .49 | .13 |
| Group | 3.53 | 1 (4) | 0.01 | .93 | .002 |
| WOLFE FMness | 19.81 | 1 (4) | 0.05 | .84 | .01 |
| PROMIS Depression | 12.23 | 1 (4) | 0.03 | .87 | .007 |
| PROMIS Anxiety | 215.68 | 1 (4) | 0.52 | .51 | .12 |
| Condition by Session | 10.73 | 1 (4) | 0.21 | .67 | .05 |
| Condition by Group | 26.00 | 1 (4) | 0.30 | .61 | .07 |
| Condition by WOLFE FMness | 25.36 | 1 (4) | 0.30 | .62 | .07 |
| Condition by PROMIS Depression | 9.75 | 1 (4) | 0.11 | .75 | .03 |
| Condition by PROMIS Anxiety | 81.60 | 1 (4) | 0.95 | .39 | .19 |
| Session by Group | 15.46 | 1 (4) | 0.03 | .88 | .007 |
| Session by WOLFE FMness | 47.75 | 1 (4) | 0.08 | .79 | .02 |
| Session by PROMIS Depression | 77.16 | 1 (4) | 0.14 | .73 | .03 |
| Session by PROMIS Anxiety | 912.78 | 1 (4) | 1.60 | .28 | .29 |
| Session by Condition by Group | 0.07 | 1 (4) | 0.001 | .97 | .000 |
| Session by Condition by WOLFE FMness | 3.16 | 1 (4) | 0.06 | .82 | .02 |
| Session by Condition by PROMIS Depression | 19.30 | 1 (4) | 0.37 | .57 | .09 |
| Session by Condition by PROMIS Anxiety | 45.17 | 1 (4) | 0.88 | .40 | .18 |

**Notes:** Adjusted heart rate variability (HRV_BL_) was assessed with repeated measures ANCOVA.

**Abbreviations:** ANCOVA, analysis of covariance; HRV_BL_, heart rate variability: root mean square of successive differences between heartbeats baseline corrected; df, degrees of freedom; FM, fibromyalgia; PROMIS, Patient-Reported Outcomes Measurement Information System.

**Supplementary Table 5** Pearson’s correlation table for Audio to Silence session change in pain and heart rate measures with fibromyalgia, depression, and anxiety symptom severity and music experience variables.

|  | WOLFE FMness  Pearson’s *r* (*p*, 2-tailed) | BPI worst 2 average | FIQR Score | PROMIS: Depression | PROMIS: Anxiety | MEQ: Commitment to Music | MEQ: Innovative Musical Aptitude | MEQ: Social Uplift | MEQ: Affective Reactions to Music | MEQ: Positive Psychotropic Effects from Music | MEQ: Reactive Musical Behavior | Temporal Summation: Audio - Silence | Mechanical Pain Tolerance: Audio - Silence | HR_BL_:  Listen - Pain; Audio - Silence | HRV_BL_:  Pain - Listen; Audio - Silence |
| --- | --- | --- | --- | --- | --- | --- | --- | --- | --- | --- | --- | --- | --- | --- | --- |
| WOLFE FMness | 1 | .75 (.02*) | .64 (.06^†^) | -.57 (.11) | -.62 (.07^†^) | .60 (.09^†^) | .15 (.69) | .01 (.98) | -.70 (.03*) | -.06 (.89) | -.24 (.53) | -.05 (.90) | .01 (.99) | .35 (.36) | .05 (.89) |
| BPI worst 2 average | .75 (.02*) | 1 | .60 (.09^†^) | -.21 (.59) | -.32 (.41) | .49 (.18) | -.03 (.94) | .06 (.87) | -.62 (.08^†^) | -.26 (.50) | -.23 (.55) | -.34 (.38) | -.33 (.39) | .15 (.71) | -.20 (.61) |
| FIQR Score | .64 (.06^†^) | .60 (.09^†^) | 1 | -.06 (.88) | -.39 (.30) | .33 (.39) | -.31 (.41) | -.03 (.94) | -.60 (.09^†^) | -.53 (.14) | -.48 (.19) | .10 (.80) | -.25 (.52) | .36 (.35) | .57 (.11) |
| PROMIS: Depression | -.57 (.11) | -.21 (.59) | -.06 (.88) | 1 | .31 (.42) | -.64 (.07^†^) | -.59 (.095^†^) | -.16 (.69) | .33 (.38) | -.54 (.13) | -.24 (.54) | .13 (.74) | -.65 (.059^†^) | .22 (.56) | .22 (.57) |
| PROMIS: Anxiety | -.62 (.07^†^) | -.32 (.41) | -.39 (.30) | .31 (.42) | 1 | -.07 (.86) | .06 (.87) | .09 (.82) | .80 (.009*) | .15 (.69) | .47 (.20) | -.65 (.057^†^) | .07 (.86) | -.78 (.01*) | -.43 (.25) |
| MEQ: Commitment to Music | .60 (.09^†^) | .49 (.18) | .33 (.39) | -.64 (.07^†^) | -.07 (.86) | 1 | .44 (.24) | -.02 (.97) | -.28 (.47) | .46 (.21) | .33 (.38) | -.36 (.34) | .18 (.65) | -.31 (.43) | -.34 (.37) |
| MEQ: Innovative Musical Aptitude | .15 (.69) | -.03 (.94) | -.31 (.41) | -.56 (.095^†^) | .06 (.87) | .44 (.24) | 1 | -.50 (.17) | .10 (.80) | .62 (.08^†^) | .86 (.003*) | -.44 (.24) | .54 (.13) | -.33 (.38) | -.34 (.37) |
| MEQ: Social Uplift | .01 (.98) | .06 (.87) | -.03 (.94) | -.16 (.69) | .09 (.82) | -.02 (.97) | -.50 (.17) | 1 | -.18 (.65) | .18 (.64) | -.46 (.22) | -.03 (.93) | .26 (.50) | -.08 (.84) | -.29 (.45) |
| MEQ: Affective Reactions to Music | -.70 (.03*) | -.62 (.08^†^) | -.60 (.09^†^) | .33 (.38) | .80 (.01*) | -.28 (.47) | .10 (.80) | -.18 (.65) | 1 | .06 (.88) | .41 (.28) | -.25 (.51) | -.07 (.86) | -.66 (.053^†^) | -.25 (.51) |
| MEQ: Positive Psychotropic Effects from Music | -.06 (.89) | -.26 (.50) | -.53 (.14) | -.54 (.13) | .15 (.69) | .46 (.21) | .62 (.08) | .18 (.64) | .06 (.88) | 1 | .66 (.054^†^) | -.30 (.44) | .71 (.03*) | -.31 (.41) | -.59 (.097^†^) |
| MEQ: Reactive Musical Behavior | -.24 (.53) | -.23 (.55) | -.48 (.19) | -.24 (.54) | .47 (.20) | .33 (.38) | .86 (.003*) | -.46 (.22) | .41 (.28) | .66 (.054^†^) | 1 | -.59 (.093^†^) | .45 (.23) | -.53 (.14) | -.46 (.21) |
| Temporal Summation: Silence - Audio | -.05 (.90) | -.34 (.38) | .10 (.80) | .13 (.74) | -.65 (.057^†^) | -.36 (.34) | -.44 (.24) | -.03 (.93) | -.25 (.51) | -.30 (.44) | -.59 (.093^†^) | 1 | -.23 (.56) | .58 (.10) | .65 (.06^†^) |
| Mechanical Pain Tolerance: Audio - Silence | .01 (.99) | -.33 (.39) | -.25 (.52) | -.65 (.059^†^) | .07 (.86) | .18 (.65) | .54 (.13) | .26 (.50) | -.07 (.86) | .71 (.03*) | .45 (.23) | -.23 (.56) | 1 | -.20 (.61) | -.13 (.75) |
| HR_BL_:  Listen - Pain; Audio - Silence | .35 (.36) | .15 (.71) | .36 (.35) | .22 (.56) | -.78 (.01*) | -.31 (.38) | -.33 (.38) | -.08 (.84) | -.66 (.05) | -.31 (.41) | -.53 (.14) | .58 (.10) | -.20 (.61) | 1 | .54 (.14) |
| HRV_BL_:  Pain - Listen;  Audio - Silence | .05 (.89) | -.20 (.61) | .57 (.11) | .22 (.57) | -.43 (.25) | -.34 (.37) | -.34 (.37) | -.29 (.45) | -.25 (.51) | -.59 (.097^†^) | -.46 (.21) | .65 (.06^†^) | -.13 (.75) | .54 (.14) | 1 |

**Notes:** All change scores were calculated so that greater analgesic response (*i.e.*, lower pain) in the audio session would be indicated by higher values. Higher values for Temporal Summation indicate worse pain; lower values for Mechanical Pain Tolerance indicate worse pain. Because heart rate was expected to decrease during pain, the change score for HR_BL_ was calculated as the between-session difference (Audio minus Silence) for Listen minus Pain. Correspondingly, vagal response as a coping mechanism was expected to increase during pain, so the change score for RRMSD_BL_ was calculated as the between-session difference (Audio minus Silence) for Pain minus Listen. *Indicates significant correlations at two-tailed *p*<.05. ^†^Indicates non-significant correlations at two-tailed *p*<.10.

**Abbreviations:** FM, fibromyalgia; BPI, Brief Pain Inventory; FIQR, Fibromyalgia Impact Questionnaire – Revised; PROMIS, Patient-Reported Outcomes Measurement Information System; MEQ, Music Experience Questionnaire; HR_BL_, hear rate baseline corrected; HRV_BL_, heart rate variability: root mean square of successive differences between heartbeats baseline corrected; df, degrees of freedom; FM, fibromyalgia; PROMIS, Patient-Reported Outcomes Measurement Information System
